# Clinical features and outcomes of 221 patients with COVID-19 in Wuhan, China

**DOI:** 10.1101/2020.03.02.20030452

**Authors:** Guqin Zhang, Chang Hu, Linjie Luo, Fang Fang, Yongfeng Chen, Jianguo Li, Zhiyong Peng, Huaqin Pan

## Abstract

**Rationale:** In late December 2019, an outbreak of acute respiratory illness, now officially named as COVID-19, or coronavirus disease 2019, emerged in Wuhan, China, now spreading across the whole country and world. More data were needed to understand the clinical characteristics of the disease.

**Objectives:** To study the epidemiology, clinical features and outcomes of patients with COVID-19.

**Methods:** we performed a single center, retrospective case series study in 221 patients with laboratory confirmed SARS-CoV-2 pneumonia at a university hospital.

**Measurements and Main Results:** The median age was 55.0 years and 48.9% were male and only 8 (3.6%) patients had a history of exposure to the Huanan Seafood Market. Compared to the non-severe pneumonia patients, the median age of the severe patients was significantly older, and they were more likely to have chronic comorbidities. Most common symptoms in severe patients were high fever, anorexia and dyspnea. On admission, 33.0% patients showed leukopenia and 73.8% showed lymphopenia. In addition, the severe patients suffered a higher rate of co-infections with bacteria or fungus and they were more likely to developing complications. As of February 15, 2020, 19.0% patients had been discharged and 5.4% patients died. 80% of severe cases received ICU care, and 52.3% of them transferred to the general wards due to relieved symptoms, and the mortality rate of severe patients in ICU was 20.5%.

**Conclusions:** The COVID-19 epidemic spreads rapidly by human-to-human transmission. Patients with elder age, chronic comorbidities, blood leukocyte/lymphocyte count, procalcitonin level, co-infection and severe complications might increase the risk of poor clinical outcomes.

## Introduction

In late December 2019, an outbreak of acute respiratory illness, now officially named as the COVID-19, or coronavirus disease 2019, emerged in Wuhan, China. ^1 2^ From bronchoalveolar lavage samples of the infected patients, a novel beta-coronavirus (SARS-CoV-2) was isolated and identified as the causative agent. ^3^ Current studies have demonstrated that the COVID-19 shares over 88% homology with two bat-derived severe acute respiratory syndrome (SARS)-related coronaviruses, bat-SL-CoVZC45 and bat-SLCoVZC21, and that bats may be the most likely natural host. ^4^ However, whether the SARS-CoV-2 transmits directly from bats or through an intermediate host is still uncertain. Although epidemiological studies indicate a common link between the initially diagnosed patients and the Huanan Seafood Market in Wuhan, ^5^ an increasing number of later confirmed cases have involved infected patients without history of contacting the implicated market, nor traveling to Wuhan, ^6^ and the family cluster type of infections were reported. ^7 8^ Human-to-human transmission has been confirmed, ^6^ and respiratory droplet and contact are the main transmission routes, while recent reports also suggested the existence of the fecal-oral transmission route. ^6 9^

Based on a recent large-scale epidemiological survey, the latency period of the SARS-CoV-2 may extend up to 24 days, even though the proportion of patients with long incubation period is very small, but the medium incubation period remains short at 3 days. ^6^ Reported illnesses have ranged from patients with little or no symptoms to patients being severely ill and dying. ^6^ The main clinical manifestations include fever, cough, fatigue, and dyspnea. ^5 10 11^ Compared to young and middle-aged patients with COVID-19, elder infected patients with chronic comorbidities have an increased risk of developing organ dysfunctions, including shock, acute respiratory distress syndrome (ARDS), acute cardiac injury and acute kidney injury, resulting in a higher mortality rate. ^10 11^ However, the clinical features between severe and non-severe cases have not yet been well described. Unlike SARS-CoV, the SARS-CoV-2 displays a highly contagious infectiousness even during the asymptomatic period ^12^. At present, there is no effective medication nor vaccine for the SARS-CoV-2 infection. As of February 22, 2020, the National Health Commission of China has reported a total of 76,936 confirmed, 4,148 suspected cases and 2442 deaths in 31 provinces across the country. ^13^ In addition, more than 2700 confirmed cases have also been reported abroad in 26 different countries.^14^

In this study, we have performed a comprehensive exploration of the epidemiological, clinical, laboratory and radiological characteristics of 221 hospitalized patients with laboratory-confirmed COVID-19, including 55 severe patients and 166 non-severe patients of Zhongnan Hospital of Wuhan University, Wuhan, China, from January 2 to February 10, 2020.

## Methods

### Patients

For this retrospective, single-center study, we recruited 221 patients who were confirmed diagnosed as COVID-19 according to WHO interim guidance, from January 2 to February 10, 2020 at Zhongnan Hospital of Wuhan University, Wuhan, China. This study was approved by the National Health Commission of China and Ethics Commission of Zhongnan Hospital. Written informed consent was waived by the Ethics Commission for emerging infectious disease.

### Procedures

221 patients were confirmed to be infected with COVID-19. The presence of SARS-CoV-2 in pharyngeal swab specimens was detected by real-time RT-PCR using ORF1ab/N gene PCR kit (Biogerm, Cat# SJ-HX-226-1,2, Shanghai, China) according to the protocol described previously. ^11^ The diagnostic criteria for RT-PCR results were based on the recommendation by the National Institute for Viral Disease Control and Prevention (China) (http://ivdc.chinacdc.cn/kyjz/202001/t20200121_211337.html). Confirmed cases were treated with effective isolation and protective conditions, and severe and critical cases were admitted to the Emergency Department or ICU wards.

All patients were monitored for vital signs (body temperature, heart rate, pulse oxygen saturation, respiratory rate and blood pressure). Initial investigations included a complete blood count, coagulation profile, and serum biochemical test (including liver and renal function, creatine kinase, lactate dehydrogenase and electrolytes). All patients were monitored by chest x-rays or CT. Respiratory specimens, including pharyngeal swabs, bronchoalveolar lavage fluid, sputum or bronchial aspirates were tested for other respiratory viruses including influenza A virus (H1N1, H7N9), influenza B virus, respiratory syncytial virus, parainfluenza virus, adenovirus, and other atypical pathogens such as mycoplasma, chlamydia and legionella pneumophila, which were also examined with real-time RT-PCR. Routine bacterial and fungal examinations were also performed by conventional microbial culture techniques.

Treatment strategies included: (1) effective oxygen therapy according to the severity of hypoxemia (nasal catheter/mask oxygen inhalation, high-flow nasal cannula (HFNC) and non-invasive ventilation (NIV)), (2) supportive treatment to ensure sufficient energy intake and balance for electrolytes and acid-base levels, (3) antiviral treatment (including oseltamivir, arbidol hydrochloride, α-interferon atomization inhalation, and lopinavir/ritonavir), (4) antibiotic therapy if accompany with bacterial infection (moxifloxacin hydrochloride; piperacillin sodium tazobactam sodium; cefoperazone sulbactam et al.), (5) intravenous corticosteroid therapy is optional and based on the clinical experiences of the doctors, (6) continuous renal replacement therapy if acute renal impairment or failure occurs. Invasive mechanical ventilation was given to patients with hypoxic respiratory failure and acute respiratory distress syndrome (ARDS). Extracorporeal membrane oxygenation (ECMO) support was used for the patients with refractory hypoxemia that is difficult to be corrected by prone and protective lung ventilation strategy.

Nucleic acid tests for SARS-CoV-2 were repeated twice and shown virus clearance before patients discharge or discontinuation of quarantine.

### Data collection

We reviewed clinical charts, nursing records, laboratory findings and chest radiography for all patients with laboratory-confirmed COVID-19 infection. The data of epidemiological, clinical, laboratory, and radiological features, treatment and outcomes were obtained from standardized data collection forms and electronic medical records. For critically ill patients admitted to the ICU, the Glasgow Coma Scale (GCS), Sequential (sepsis-related) Organ Failure Assessment (SOFA), and Acute Physiology and Chronic Health Evaluation II (APACHE II) scores were monitored on the day of ICU admission. Two researchers independently reviewed the data to double check the correct of collected data. For the data which were not available for electronic medical records, the researchers directly communicated with patients and doctors to ascertain the data integrity. The clinical outcomes (i.e. discharges, mortality, hospitalization) were followed up to February 15, 2020.

### Definitions

The diagnostic criteria of ARDS were based on the 2012 Berlin definition. ^15^ Shock were defined according to the interim guidance of WHO for novel coronavirus-China. Hypoxemia was defined as partial arterial oxygen pressure (PaO_2_) over inspiratory oxygen fraction (FiO_2_) of less than 300 mmHg. Acute kidney injury (AKI) was identified and classified by the highest serum creatinine level or urine output criteria according to the kidney disease improving global outcomes classification. Cardiac injury was diagnosed if serum levels of cardiac biomarkers (i.e. hypersensitive troponin I) were above the 99^th^ percentile upper reference limit, or newly onset abnormalities shown in electrocardiogram or cardiac ultrasound. The severe or critical COVID-19 patients were defined according to the following criteria: fever plus one of these conditions, including respiratory rate ≥ 30 breaths/min, severe respiratory distress, SpO2 ≤ 93% on room air, occurrence of respiratory failure requiring mechanical ventilation, shock and other organ failure. ^8^

### Statistical analysis

All the continuous variables were determined the normality of the distribution by Kolmogorov-Smirnov test, the normally distributed variables were described as the means ± standard deviation (SD) and the skewed distributed variables were expressed as the median and interquartile range (IQR). We compared the normally distributed continuous variables by using the *Student t-test* and skewed distributed variables by using the *Mann-Whitney U test*. Comparisons of categorical variables between groups were conducted using *Pearson’s chi-squared test* or *Fisher’s exact test*, as appropriate. All the statistical analyses were performed using the IBM SPSS version 24.0 (IBM Corp., Armonk, NY, USA). *P* values less than 0.05 represented statistical significance and all reported *P* values were two-sided.

## Results

### Epidemiological and Clinical characteristics

The study population included a total of 221 admitted patients with confirmed COVID-19 infection in Zhongnan Hospital of Wuhan University, between January 2, 2019 and February 10, 2020. The median age was 55.0 years (IQR, 39.0-66.5; range, 20-96 years), and 108 of 221 (48.9%) were male. The numbers of patients with COVID-19 below the age of 45 years, between 45-65 years and above 65 years were 73 (33.0%), 86 (38.9%) and 62 (28.1%), respectively. Of these patients, 55 (24.9%) were severe patients and 166 (75.1%) patients were non-severe (**Table 1**). Of the whole study population, only 8 (3.6%) patients with COVID-19 had a history of exposure to the Huanan Seafood Market, and 19 (8.6%) patients suffered a secondary infection during hospitalization (**Table 1**).

**Table 1:** Demographics and baseline characteristics of patients with COVID-19

| Characteristics | Total<br>(n=221) | Severe (n=55) | Non-severe<br>(n=166) | P Value |
| --- | --- | --- | --- | --- |
| Age, years | 55.0(39.0-66.5) | 62.0(52.0-74.0) | 51.0(36.0-64.3) | <0.001 |
| <45 | 73(33.0) | 6(10.9) | 67(40.4) | <0.001 |
| 45-65 | 86(38.9) | 25(45.5) | 61(36.7) | 0.251 |
| >65 | 62(28.1) | 24(43.6) | 38(22.9) | 0.003 |
| Sex |  |  |  | 0.011 |
| Male | 108(48.9) | 35(63.6) | 73(44.0) |  |
| Female | 113(51.1) | 20(36.4) | 93(56.0) |  |
| Huanan Seafood Market<br>exposure | 8(3.6) | 3(5.5) | 5(3.0) | 0.414 |
| Infection during hospitalization | 19(8.6) | 9(16.4) | 10(6.0) | 0.026 |
| **Comorbidity** | 78(35.3) | 40(72.7) | 38(22.9) | <0.001 |
| Hypertension | 54(24.4) | 26(47.3) | 28(16.9) | <0.001 |
| Diabetes | 22(10.0) | 7(12.7) | 15(9.0) | 0.428 |
| Cardiovascular disease | 22(10.0) | 13(23.6) | 9(5.4) | <0.001 |
| Cerebrovascular disease | 15(6.8) | 11(20.0) | 4(2.4) | <0.001 |
| COPD | 6(2.7) | 4(7.3) | 2(1.2) | 0.035 |
| CKD | 6(2.7) | 5(9.1) | 1(0.6) | 0.004 |
| Chronic liver disease | 7(3.2) | 4(7.3) | 3(1.8) | 0.066 |
| Malignancy | 9(4.1) | 4(7.3) | 5(3.0) | 0.231 |
| Immunosuppression | 3(1.4) | 1(1.8) | 2(1.2) | 1.000 |
| **Signs and symptoms** |  |  |  |  |
| Fever | 200(90.5) | 55(100.0) | 145(87.3) | 0.006 |
| Highest temperature °C | 38.5(38.0-39.0) | 38.8(38.5-39.0) | 38.3(37.7-38.9) | <0.001 |
| <37.3 °C | 21(9.5) | 0(0) | 21(12.7) | 0.006 |
| 37.3-38.0°C | 57(25.8) | 6(10.9) | 51(30.7) | 0.004 |
| 38.1-39.0°C | 104(47.1) | 36(65.5) | 68(41.0) | 0.002 |
| >39.0°C | 39(17.6) | 13(23.6) | 26(15.7) | 0.179 |
| Fatigue | 156(70.6) | 42(76.4) | 114(68.7) | 0.278 |
| Cough | 136(61.5) | 36(65.5) | 100(60.2) | 0.491 |
| Anorexia | 80(36.2) | 34(61.8) | 46(27.7) | <0.001 |
| Dyspnea | 64(29.0) | 35(63.6) | 29(17.5) | <0.001 |
| Diarrhea | 25(11.3) | 9(16.4) | 16(9.6) | 0.172 |
| Pharyngalgia | 22(10.0) | 8(14.5) | 14(8.4) | 0.189 |
| Headache | 17(7.7) | 4(7.3) | 13(7.8) | 1.000 |
| Abdominal pain | 5(2.3) | 2(3.6) | 3(1.8) | 0.429 |
| Onset of symptom to hospital admission (d) | 7.0(4.0-10.0) | 7.0(6.0-10.0) | 7.0(4.0-9.0) | 0.041 |
| Onset of symptom to dyspnea (d) | 8.0(4.0-11.0) | 8.0(5.0-11.0) | 5.0(2.0-7.8) | 0.006 |
| Heart rate (beats/minute) | 84(76-96) | 88(80-105) | 81(76-95) | <0.01 |
| Respiratory rate | 20(19-21) | 21(20-26) | 20(19-20) | <0.001 |
| Mean arterial pressure (mmHg) | 90(84-97) | 92(80-97) | 89(84-96) | 0.930 |

Data are median (IQR), n (%). *P* values comparing severe and non-severe are from $\chi^2$ test, Fisher's exact test, or Mann-Whitney U test. COPD = chronic obstructive pulmonary disease. CKD = chronic kidney disease. COVID-19 = Corona Virus Disease 2019.

In total, 78 (35.3%) patients had 1 or more chronic comorbidities, including hypertension (54 [24.4%]), diabetes (22 [10.0%]), cardiovascular disease (22 [10.0%]), cerebrovascular disease (15 [6.8%]), chronic obstructive pulmonary disease (COPD) (6 [2.7%]), chronic kidney disease (CKD) (6 [2.7%]), chronic liver disease (7 [3.2%]), malignancy (9 [4.1%]) and patients with immunosuppression treatment (3 [1.4%]) (**Table 1**).

The symptom onset date of the first patient identified was Dec 25, 2019. The most common symptoms were fever (200 [90.5%]), followed by fatigue (156 [70.6%]), cough (136 [61.5%]), anorexia (80 [36.2%]) and dyspnea (64 [29.0%]). Less common symptoms included diarrhea (25 [11.3%]), pharynglagia (22 [10.0%]), headache (17 [7.7%]) and abdominal pain (5 [2.3%]). The median duration from onset of symptoms to hospital admission was 7.0 days (IQR, 4.0-10.0), to dyspnea was 8.0 days (IQR, 4.0-11.0), and to ICU admission was 10.0 days (IQR, 7.0-13.0) (**Table 1** and **3**).

Compared to the non-severe patients (n = 166), the median age of the severe patients was significantly older (62.0 years [IQR, 52.0-74.0] vs 51.0 years [IQR, 36.0-64.3]; *P* < 0.001). 35 (63.6%) of the severe patients were male, and 93 (56.0%) of the non-severe patients were female. The severe patients were also more likely to have underlying comorbidities (40 [72.7%] vs 38 [22.9%]; *P* < 0.001), including hypertension (26 [47.3%] vs 28 [16.9%]; *P* < 0.001), cardiovascular disease (13 [23.6%] vs 9 [5.4%]; *P* < 0.001) and cerebrovascular disease (11 [20.0%] vs 4 [2.4%]; P < 0.001) (**Table 1**). A higher proportion of severe patients developed symptoms such as high fever (body temperature above 38.1 °C, 49 [89.1%] vs 94 [56.6%]; *P* < 0.001), anorexia (34 [61.8%] vs 46 [27.7%]; *P* < 0.001) and dyspnea (35 [63.6%] vs 29 [17.5%]; *P* < 0.001) (**Table 1**). Vital signs including heart rate (88 [80-107] vs 80 [74-90]; *P* < 0.01) and respiratory rate (21 [20-26] vs 20 [19-20]; *P* < 0.001) were also significantly increased in severe patients compared to non-severe patients (**Table 1**).

### Laboratory findings in severe and non-severe patients

On admission, the blood counts of 73 (33.0%) of the 221 patients showed leukopenia (white blood cell count < 3.5×10^9^/L) and 163 (73.8%) showed lymphopenia (lymphocyte count < 1.1×10^9^/L). There were numerous laboratory parameters were significantly increased in severe patients (**Table 2**), including the white blood cell and neutrophil, the prothrombin time, levels of D-dimer, hypersensitive troponin I, creatine kinase, creatine kinase-MB, lactate dehydrogenase, alanine and aspartate aminotransferase (ALT/AST), total bilirubin, serum creatinine as well as procalcitonin (**Table 2**, *P* < 0.001). Additionally, the lymphocyte count was significantly decreased in severe patients compared to the non-severe patients (**Table 2**, *P* < 0.001)). Of the 221 patients with COVID-19, a total of 215 (97.3%) showed bilateral and the rest (6, [2.7%]) showed unilateral chest radiographs abnormalities, characterized by multiple patchy, ground-grass opacities and multiple lobes of consolidation (the 1^st^ and 2^nd^ chest CT in **Figure 1A**) or honeycomb-like thickening of the interlobular septa and sub-segmental areas of consolidation (the 1^st^ and 2^nd^ chest CT in **Figure 1B**). Pathogenic analyses show that the severe patients suffered a significantly higher rate of co-infections with bacteria (14 [25.5%] vs 3 [1.8%]; *P* < 0.001) and fungus (6 [10.9%] vs 1 [0.6%]; *P* = 0.001) compared to the non-severe patients (**Table 2**). Of the 55 severe COVID-19 patients, 44 (80%) of them were admitted to the ICU due to combined moderate or severe ARDS, requiring non-invasive or invasive mechanical ventilation therapy. The median time from onset of symptoms to ICU admission was 10.0 days (IQR, 7-13) (**Table 3**). On the day of ICU admission, the median GCS, APACHE II, and SOFA scores were 15 (IQR, 11-15), 17 (IQR, 12-22) and 5 (IQR, 4-8), respectively (**Table 3**), indicating critical illness. The median PaO_2_ level of patients in ICU was 66 mm/Hg (IQR, 53-87) and the median of P/F (PaO_2_ to FiO_2_) ratio was 113 mm/Hg (IQR, 84-190).

**Table 2:** Laboratory tests of patients with COVID-19 on admission to hospital

| Laboratory parameters | Total<br>(n=221) | Severe<br>(n=55) | Non-severe<br>(n=166) | P Value |
| --- | --- | --- | --- | --- |
| White blood cell count ($\times 10^9$ /L; normal range 3.5–9.5) | 4.4(3.2-6.6) | 6.2(4.1-9.4) | 4.1(3.1-5.8) | <0.001 |
| <3.5 | 73(33.0) | 11(20.0) | 62(37.3) | 0.018 |
| 3.5-9.5 | 125(56.6) | 31(56.4) | 94(56.6) | 0.973 |
| >9.5 | 23(10.4) | 13(23.6) | 10(6.0) | <0.001 |
| Neutrophil count ($\times 10^9$ /L; normal range 1.8-6.3) | 3.0(1.9-5.1) | 5.4(2.8-8.4) | 2.6(1.8-4.0) | <0.001 |
| Lymphocyte count ($\times 10^9$ /L; normal range 1.1-3.2) | 0.8(0.6-1.1) | 0.7(0.4-0.9) | 0.9(0.6-1.2) | <0.001 |
| <0.5 | 39(17.6) | 18(32.7) | 21(12.7) | 0.001 |
| 0.5-1.1 | 124(56.2) | 30(54.5) | 94(56.6) | 0.788 |
| >1.1 | 58(26.2) | 7(12.7) | 51(30.7) | 0.009 |
| Monocyte count ($\times 10^9$ /L; normal range 0.1-0.6) | 0.4(0.3-0.5) | 0.4(0.2-0.5) | 0.4(0.3-0.5) | 0.381 |
| Platelet count ($\times 10^9$ /L; normal range 125-350) | 175(127-209) | 169(111-202) | 175(136-213) | 0.050 |
| Prothrombin time (s; normal range 9.4-12.5) | 12.9(12.1-13.6) | 13.4(12.3-14.8) | 12.7(12.1-13.4) | <0.001 |
| Activated partial thromboplastin time (s; normal range 25.1-36.5) | 31.1(29.1-33.1) | 31.1(29.0-34.9) | 31.1(29.1-33.0) | 0.666 |
| D-dimer (mg/L; normal range 0-500) | 227(129-485) | 443(211-1304) | 184(118-324) | <0.001 |
| Hypersensitive troponin I (pg/mL; normal range <26.2) | 7.6(3.6-21.5) | 14.9(6.9-55.3) | 5.4(2.2-9.7) | <0.001 |
| Creatine kinase (U/L; normal range <171) | 87(55-143) | 121(73-268) | 75(53-122) | <0.001 |
| Creatine kinase-MB (U/L; normal range <25) | 13(10-17) | 18(14-35) | 12(10-15) | <0.001 |
| Lactate dehydrogenase (U/L; normal range 125-243) | 227(174-367) | 424(287-591) | 204(167-290) | <0.001 |
| Alanine aminotransferase (U/L; normal range 9-50) | 23(16-39) | 32(22-57) | 22(14-33) | <0.001 |
| Aspartate aminotransferase (U/L; normal range 15-40) | 29(22-49) | 51(29-78) | 27(20-38) | <0.001 |
| Total bilirubin (mmol/L; normal range 5-21) | 10.0(8.0-14.2) | 11.4(8.6-17.4) | 9.6(7.9-13.8) | 0.034 |
| Blood urea nitrogen (mmol/l; normal range 2.8-7.6) | 4.3(3.4-5.6) | 5.8(4.3-8.5) | 4.0(3.3-5.0) | <0.001 |
| Creatinine (μmol/L; normal range 64-104) | 69(56-84) | 75(64-108) | 67(55-79) | <0.001 |
| Procalcitonin (ng/mL; normal range <0.05) |  |  |  |  |
| <0.05 | 150(67.9) | 15(27.3) | 135(83.1) | <0.001 |
| 0.05-1.00 | 58(26.2) | 28(50.9) | 30(18.1) | <0.001 |
| >1.00 | 13(5.9) | 12(21.8) | 1(0.6) | <0.001 |
| Bilateral involvement of chest radiographs | 215(97.3) | 55(100.0) | 160(96.4) | 0.340 |
| **Co-infection** |  |  |  |  |
| Other viruses | 33(14.9) | 16(29.1) | 17(10.2) | 0.001 |
| Bacteria | 17(7.7) | 14(25.5) | 3(1.8) | <0.001 |
| Fungus | 7(3.2) | 6(10.9) | 1(0.6) | 0.001 |

Data are median (IQR), n (%). *P* values comparing severe and non-severe are from $\chi^2$ test, Fisher's exact test, or Mann-Whitney U test. COVID-19 = Corona Virus Disease 2019.

**Table 3:** The scoring system and blood gas analysis of patients with COVID-19 on admission to ICU

|  | Normal range | ICU (n=44) |
| --- | --- | --- |
| Onset of symptom to ICU admission (d) | NA | 10.0(7.0-13.0) |
| GCS | NA | 15(11-15) |
| APACHE II | NA | 17(12-22) |
| SOFA | NA | 5(4-8) |
| PH | 7.35-7.45 | 7.41(7.39-7.47) |
| Lactate, mmol/l | 0.5-1.6 | 1.5(0.8-2.2) |
| PaO<sub>2</sub>, mm/Hg | 83-108 | 66(53-87) |
| PaO<sub>2</sub>:FiO<sub>2</sub>, mm/Hg | 400-500 | 113(84-190) |
| PaCO<sub>2</sub>, mm/Hg | 35-48 | 36(32-40) |

Data are median (IQR). GCS = Glasgow Coma Score; APACHE II = Acute Physiology and Chronic Health Evaluation II; SOFA = Sequential Organ Failure Assessment; PaO<sub>2</sub> = Partial Pressure of Oxygen; PaCO<sub>2</sub> = Partial Pressure of Carbon Dioxide; FiO<sub>2</sub> = fraction of inspiratory oxygen. ICU = intensive care unit; NA, not available.

**Figure 1:**
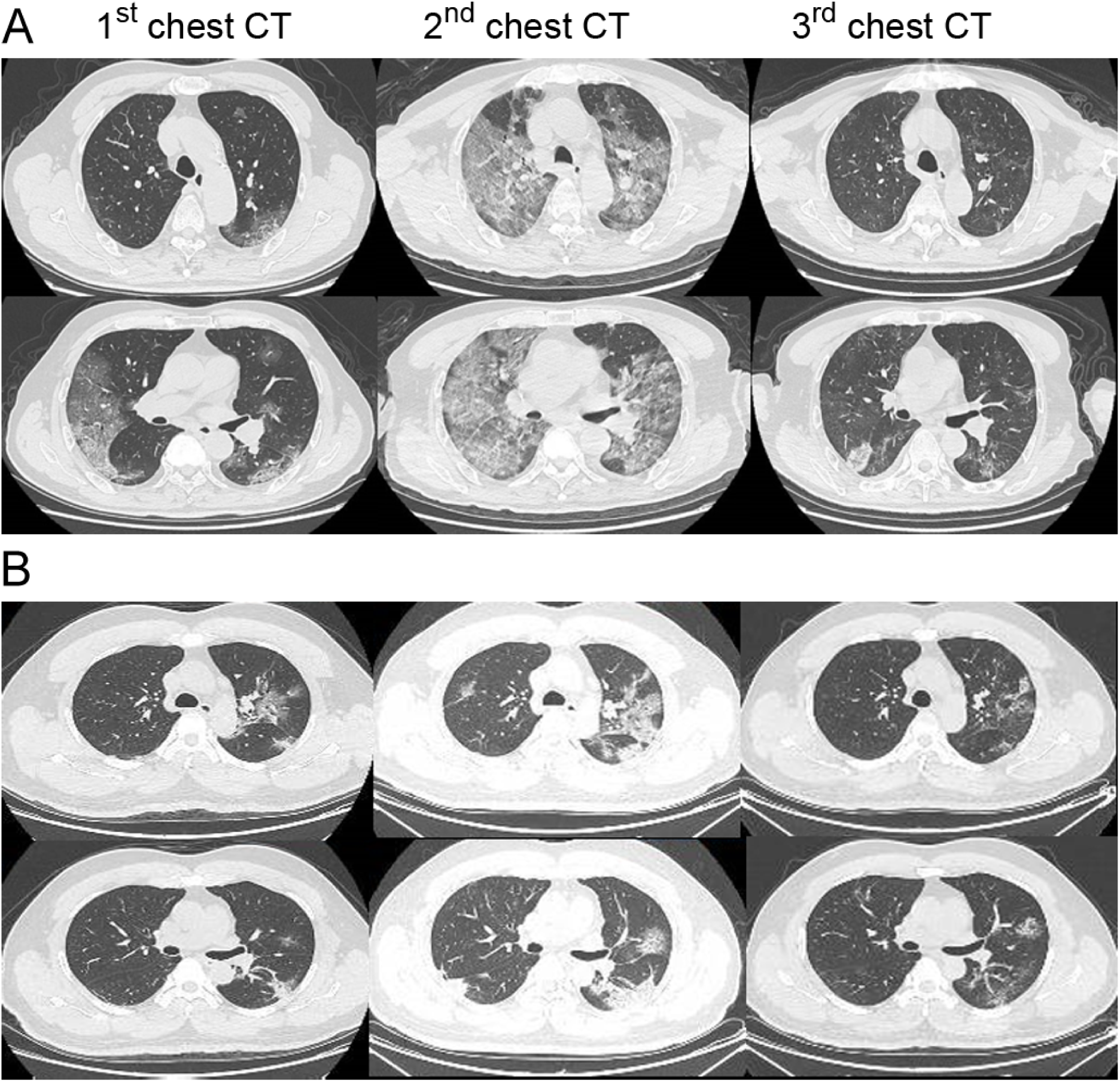
Transverse chest CT images of the patients with COVID-19. **Case A**:Transverse chest CT images from a 72-year-old man with severe COVID-19. The 1^st^ chest CT shows multiple ground-glass opacities in bilateral lungs on day 8 after symptom onset. The 2^ed^ chest CT Shows progressive bilateral ground-glass opacities and multiple lobes of consolidation on day 14 after symptom onset, and the 3^ed^ images were obtained after ECMO supportive therapy in the ICU showing recovery status on day 30 after symptom onset. **Case B:** Transverse chest CT images from a 44-year-old man with mild COVID-19. The 1^st^ chest CT shows bilateral multiple lobular and subsegmental areas of consolidation on day 7 after symptom onset. The 2^ed^ chest CT Shows bilateral ground-glass opacity and subsegmental areas of consolidation on day 10 after symptom onset, and the 3^ed^ chest CT shows improved status on day 18 after symptom onset.

### Complications, Treatment and Prognosis

Common complications among the total 221 subjects included ARDS, arrhythmia, acute cardiac injury, acute kidney injury (AKI) and shock. Compared to non-severe patients, the percentages of severe patients with complications were significantly increased, including ARDS (48 [87.3%] vs 0; *P* < 0.001), arrhythmia (22 [40.0%] vs 2 [1.2%]; *P* < 0.001), acute cardiac injury (16 [29.1%] vs 1 [0.6%]; *P* < 0.001), shock (15 [27.3%]) vs 0; *P* < 0.001) and AKI (8 [14.5%] vs 2 [1.2%]; *P* < 0.001) (**Table 4**).

**Table 4:** Treatments and prognosis of patients with COVID-19

| Treatments and Prognosis | Total (n=221) | Severe (n=55) | Non-severe (n=166) | P Value |
| --- | --- | --- | --- | --- |
| **Complications** |  |  |  |  |
| ARDS | 48(21.7) | 48(87.3) | 0(0) | <0.001 |
| Arrhythmia | 24(10.9) | 22(40.0) | 2(1.2) | <0.001 |
| Acute cardiac injury | 17(7.7) | 16(29.1) | 1(0.6) | <0.001 |
| Shock | 15(6.8) | 15(27.3) | 0(0) | <0.001 |
| AKI | 10(4.5) | 8(14.5) | 2(1.2) | <0.001 |
| **Treatment** |  |  |  |  |
| Antiviral therapy | 196(88.7) | 50(90.9) | 146(88.0) | <0.001 |
| Glucocorticoid therapy | 115(52.0) | 40(72.7) | 75(45.2) | <0.001 |
| CRRT | 5(2.3) | 4(7.3) | 1(0.6) | 0.016 |
| NIV | 27(12.2) | 25(45.5) | 2(1.2) | <0.001 |
| IMV | 16(7.2) | 16(29.1) | 0(0) | <0.001 |
| IMV+ECMO | 10(4.5) | 10(18.2) | 0(0) | <0.001 |
| **Prognosis** |  |  |  |  |
| Hospitalization | 168(76.0) | 37(67.3) | 131(78.9) | 0.080 |
| Discharge | 42(19.0) | 7(12.7) | 35(21.1) | 0.171 |
| Death | 12(5.4) | 12(21.8) | 0(0) | <0.001 |

Data are n (%). *P* values comparing severe and non-severe are from $\chi^2$ test, Fisher's exact test. ARDS= acute respiratory distress syndrome. AKI = acute kidney injury.
CRRT = continuous renal replacement therapy; NIV = non-invasive ventilation; IMV = invasive mechanical ventilation ECMO = extracorporeal membrane oxygenation.
COVID-19 = Corona Virus Disease 2019.

Most patients (196 [88.7%]) received antiviral therapy, and a total of 64 (49.6%) patients were given glucocorticoid treatment. The severe patients receiving antiviral therapy (50 [90.0%] vs 146 [88.0%]; *P* < 0.001) and glucocorticoid treatment (40(72.7%) vs 75 [45.2%]; *P* < 0.001) were significantly higher than those of the non-severe patients (**Table 4**).

Among all the severe patients, 25 of patients (45.5%) received non-invasive ventilation (NIV) and 26 patients (47.3%) received IMV, which were significantly higher than those of non-severe patients, respectively (45.5% vs 1.2% and 47.3% vs 0; *P* < 0.001) (**Table 4**). In addition, 48 (87.3%) of severe patients developed ARDS, and 10 (18.2%) of them were treated with IMV plus ECMO support and 2 AKI patients underwent CRRT. Of 10 severe patients receiving IMV+ECMO support, 2 patients had clinical benefits and had been discharged and 3 of them were non-survivors. The rest 5 patients were still under treatment at the time of data collection. **Figure 1A** represents the chest CT was significantly improved after receiving IMV+ECMO support.

We also analyzed the outcome of the 44 severe patients in the ICU (**Table 5**). Of these patients, 23 of them (52.3%) were symptomatic relief and transferred to the general wards, while 9 of them (20.5%) were dead, and the rest of the patients were still under treatment. The patients with higher scores of APACHE II, SOFA and increased levels of PCT, showed a worse prognosis (**Table 5**). The elder and male patients had an increased mortality rate. The dose and duration of intravenous glucocorticoid treatment showed no difference in outcomes of symptomatic relief and death (**Table 5**). Intriguingly, the patients in the death group received a significantly enlarged volume of the fluid balance per day (483 [IQR, 333∼717] vs 60 [IQR, -164∼133]; *P* < 0.001), and cumulative fluid volume in total (4800 [IQR, 2500-8996] vs 270 [IQR, -1150∼1200]; *P* < 0.001), compared to the patients in the ICU-to-Ward transfer group.

**Table 5.** Comparison of clinical parameters based on ICU outcomes: Death vs Transfer

| Parameters | ICU-to-Ward<br>Transfers<br>(n=23) | Death in ICU<br>(n=9) | P Value |
| --- | --- | --- | --- |
| Age, years | 62.0(49.0-71.0) | 76.0(57.5-81.5) | 0.093 |
| Sex |  |  | 0.681 |
| Male | 15(65.2) | 7(77.8) |  |
| Female | 8(34.8) | 2(22.2) |  |
| APACHE II on ICU admission | 13.0(10.0-16.0) | 19.0(15.0-31.5) | 0.003 |
| SOFA on ICU admission | 4.0(3.0-5.0) | 7.0(4.0-11.0) | 0.009 |
| PCT<sub>max</sub> (ng/ml) | 0.17(0.05-1.06) | 1.89(1.53-8.67) | 0.001 |
| **Co-infection** |  |  |  |
| Other viruses | 2(8.7) | 4(44.4) | 0.038 |
| Bacteria | 6(26.1) | 5(55.6) | 0.213 |
| Fungus | 3(13.0) | 4(44.4) | 0.076 |
| Onset of symptom to dyspnea (d) | 10.0(7.0-12.0) | 10.0(8.0-10.5) | 0.914 |
| Onset of symptom to ICU admission (d) | 10.0(7.0-12.0) | 11.0(8.0-12.0) | 0.832 |
| Onset of symptom to IMV (d) | 11.0(9.3-14.3) | 11.0(9.0-13.5) | 0.646 |
| Onset of symptom to glucocorticoid therapy (d) | 9.5(7.0-11.5) | 11.0(8.5-15.5) | 0.206 |
| Maximum methylprednisolone dosage (mg/d) | 80(60-80) | 80(50-100) | 0.856 |
| Duration of glucocorticoid therapy (d) | 6.5(4.0-12.0) | 8.0(2.5-12.0) | 0.776 |
| Cumulative fluid balance in ICU (ml) | 270(-1150~1200) | 4800(2500~8996) | <0.001 |
| Mean daily fluid balance in ICU (ml/d) | 60(-164~133) | 483(333~717) | <0.001 |
| ICU length of stay (d) | 8.0(5.0-13.0) | 11.0(4.5-14.5) | 0.737 |

Data are median (IQR), n (%). P values comparing severe and non-severe are from $\chi^2$ test, Fisher's exact test, or Mann-Whitney U test. COVID-19 = Corona Virus Disease 2019. ICU = intensive care unit; APACHE II = Acute Physiology and Chronic Health Evaluation II score; SOFA = Sequential Organ Failure Assessment score; PCT = procalcitonin; Cumulative fluid balance = total fluid input minus output in ICU;

As of Feb 15, 2020, a total of 42 (19.0%) patients had been discharged and 12 (5.4%) patients had died, and 168 (76.0%) patients were still hospitalized. Of the 55 severe patients, 37 (67.3%) were still hospitalized, and 7 (12.7%) had been discharged and 12 (21.8%) were dead. The mortality rate was significantly higher in the severe patients compared to that in the non-severe patients (12 [21.8%] vs 0 [0.0%]; *P* < 0.001, **Table 4**).

## Discussion

This retrospective, single-center study included a total of 221 SARS-CoV-2 pneumonia cases from Jan 2 to Feb 10, 2020. Only 8 (3.6%) of patients had a history of exposure to the Huanan Seafood Market, while a majority of patients without the exposure history indicate the rapid human-to-human transmission. Based on a recent review, the estimated mean R_0_, an indicator of virus transmissibility, for the SARS-CoV-2 is around 3.28, which is higher than the WHO estimated at 1.95.^16^, and is also higher than that of the SARS-CoV outbreak in 2003, with R_0_ approximately at 3.0.^17^

Clinical characteristic analysis shows a significantly increased age as well as elevated numbers of underlying comorbidities in severe patients than those in non-severe patients, indicating that the age and comorbidity may be important risk factors for poor outcome. Contradictory to the previous report showing a higher incidence of COVID-19 in male patients, ^5^ recent data including ours, consistently showed a similar proportion of male and female patients with COVID-19. ^11^

The patients with COVID-19 also demonstrated a decreased lymphatic count comparing to healthy people, and it was significantly decreased in severe patients compared to those in non-severe patients. Our data show that lymphocytopenia occurred in more than 80% of severe patients, consistent with the result of a recent cohort study. ^18^ Lymphocytopenia is also a prominent feature of severe patients with SARS-CoV and MERS infection which attribute to the necrosis or apoptosis of lymphocyte caused by the invasive viral particles. ^19 20^ We also analyzed the absolute number of different subsets of lymphocytes in the ICU patients. Compared to the health population, the total numbers of T cells (312 [231-558] vs [805-4459]; number per μl), CD4^+^ T cells (245 [148-297] vs [345-2350]), CD8^+^ T cells (81 [64-230] vs [345-2350]), total B cells (81 [47-106] vs [240-1317]) as well as natural killer (NK) cells (51 [21-115] vs [210-1514]) were dramatically decreased in the severe patients (data not shown), suggesting that the severity of lymphocytopenia reflects the severity of COVID-19. A recent study showed a significantly reduced total number of T cells, CD4^+^ and CD8^+^ T cells in elder and severe patients with COVID-19 ^21^, and revealed one possible mechanism by which the aberrant cytokine signaling including TNF-a, IL-6 and IL-10 may be mediated in T cell pro-apoptosis. ^21^ In addition, abnormal laboratory findings in the severe patients also included prolonged thrombin time, increased AST/AST, hypersensitive troponin I and serum creatine, suggesting aberrant coagulation pathway, hepatic injury, myocardial injury, and kidney injury, respectively. The levels of PCT, a marker suggesting the bacterial infection, were not elevated in the patients with COVID-19, suggesting the viral-mediated pneumonia rather than bacteria. These laboratory abnormalities are similar to those previously observed in patients with MERS-CoV ^22^ and SARS-CoV infection. ^23 24^

The rates of co-infection, including other viruses, bacteria and fungus, were significantly increased in severe patients with COVID-19 than those in non-severe patients. The co-infection rate was also higher in the death group than that of the ICU-to-Ward transfer group, despite no significantly statistical difference due to a small number of cases involved in. The main reasons for the increased hospital acquired infections were due to lymphocytopenia and the reduced host immune functions in critically ill patients. These patients bear invasive catheters including endotracheal tube, arteriovenous catheters, urinary and gastric tubes, resulting in an increased susceptibility to secondary infections of nosocomial multidrug-resistant pathogens, such as *A. baumannii, Escherichia coli, Pseudomonas aeruginosa* and *Enterococcus*. We found that 5 (55.6%) of COVID-19 patients co-infected with *carbapenem-resistant A. baumanni* (CRAB) in the ICU death group, which was much higher than those (4 [17.4%]) in the ICU-to-Ward transfer group (data not shown). The higher rate of CRAB infection makes difficulties in antibiotic treatment, resulting in increased possibility to develop septic shock and death. ^25^

At present, there is no evidence showing specific drug treatment against the new coronavirus in suspected or confirmed cases. The principles of treatment include improvement of the symptoms and underlying diseases, active prevention of potential complications and secondary infections. Like other viruses, the SARS-CoV-2 enters cells through receptor-mediated endocytosis.^26^ Studies showed that the SARS-CoV-2 may infect alveolar epithelial cells in lung through the angiotensin-converting enzyme II (ACE2) receptor, which is also expressed in other tissues, such as kidney, blood vessels and heart.^26 27^ Researchers using an artificial intelligence predicted that Baricitinib, an inhibitor of AP2-associated protein kinase 1 (AAK1), might be useful to interrupt the entrance of virus to cells and also the intracellular assembly of the virus particles. ^4 28^ In addition, a case report showed that remdesivir, an adenosine analogue, has shown survival benefits in one severe COVID-19 pneumonia patient ^29^. The effectiveness has been verified in vitro. ^30^ Now a couple of clinical trials focus on the efficacy of remdesivir, as well as other therapeutic strategies, such as immunoglobulins, Vitamin C infusion, mesenchymal stem cell treatment, arbidol hydrochloride plus interferon atomization, ritonavir combined with oseltamivir, lopinavir plus ritonavir and arbidol hydroxychloroquine and methylprednisolone. ^31^

The corticosteroid therapy regarding the onset therapeutic time, the dosage, and duration were still controversial in the treatment of severe SARS or SARS-CoV-2 pneumonia. ^32^ We treated corticosteroid to the patients with refractory high fever, exacerbation of wheezing symptoms, increased interstitial exudation based on chest radiology, and high levels of inflammatory mediators to inhibit the “cytokine storm”. Corticosteroid therapy (methylprednisolone 1-2 mg/kg/day) are recommended for severely ill patients with ARDS, for as short a duration of treatment as possible. ^33 34^ The results showed that the early onset use of corticosteroid may have clinical benefits, but more cases and multivariate correlation analysis regarding the safety and efficacy were needed to verify. Our data also suggested that excessive fluid resuscitation may increase the risk of death. One principle of the ARDS treatments is the restricted fluid resuscitation strategy to prevent exacerbation of pulmonary edema. ^35^

The limitation of this study is that among the 221 cases, most of the patients were still hospitalized at the time of data collected. It is incomplete to assess risk factors for outcomes. Continued observation and dynamic clinical dataset are required.

## Conclusion

In this single-center case series of 221 hospitalized patients with confirmed COVID-19 in Wuhan, China, 55 severe patients (24.9%) with elder age and chronic comorbidities, developed more than one complication. 44 (80%) of them received ICU care, and 52.3% of them transferred to the general wards due to relieved symptoms, and the mortality rate of severe patients in ICU was 20.5%. Of the 166 non-severe patients, 21.1% of them were cured and discharged and no patients died. Older and male patients with higher APACHE II & SOFA scores, elevated PCT level, excessive fluid volume input, as well as the delayed use of corticosteroid might increase the risk of death.

## Data Availability

We declare all the data referred to our manuscript are available.

## Acknowledgments

This work was supported by the National Natural Science Foundation of China (grant no. 81700493 to Dr. Pan; grant no. 81971816 to Dr. Peng).

## Ethical approval

This study was approved by the National Health Commission of China and Ethics Commission of Zhongnan Hospital.

## References

1. Lu H, Stratton CW, Tang YW. Outbreak of Pneumonia of Unknown Etiology in Wuhan China: the Mystery and the Miracle. Journal of Medical Virology 2020 Apr;92(4):401–402. doi: 10.1002/jmv.25678.

2. Centers for Disease Control and Prevention (CDC): Coronavirus Disease 2019 (COVID-19) Situation Summary. https://wwwcdcgov/coronavirus/2019-ncov/summaryhtml

3. Zhu N, Zhang D, Wang W, et al. A novel coronavirus from patients with pneumonia in China, 2019. New England Journal of Medicine 2020. Feb 20;382(8):727–733. doi: 10.1056/NEJMoa2001017.

4. Lu R, Zhao X, Li J, et al. Genomic characterisation and epidemiology of 2019 novel coronavirus: implications for virus origins and receptor binding. The Lancet 2020. Feb 22;395(10224):565–574. doi: 10.1016/S0140-6736(20)30251-8.

5. Chen N, Zhou M, Dong X, et al. Epidemiological and clinical characteristics of 99 cases of 2019 novel coronavirus pneumonia in Wuhan, China: a descriptive study. The Lancet 2020. Feb 15;395(10223):507–513. doi: 10.1016/S0140-6736(20)30211-7.

6. Guan W-j, Ni Z-y, Hu Y, et al. Clinical characteristics of 2019 novel coronavirus infection in China. medRxiv 2020:2020.02.06.20020974. doi: 10.1101/2020.02.06.20020974.

7. Chan JF-W, Yuan S, Kok K-H, et al. A familial cluster of pneumonia associated with the 2019 novel coronavirus indicating person-to-person transmission: a study of a family cluster. The Lancet 2020. Feb 15;395(10223):514–523. doi: 10.1016/S0140-6736(20)30154-9.

8. Jin Y-H, Cai L, Cheng Z-S, et al. A rapid advice guideline for the diagnosis and treatment of 2019 novel coronavirus (2019-nCoV) infected pneumonia (standard version). Military Medical Research 2020;7(1):4. doi: 10.1186/s40779-020-0233-6.

9. Zhang H, Kang Z, Gong H, et al. The digestive system is a potential route of 2019-nCov infection: a bioinformatics analysis based on single-cell transcriptomes. bioRxiv 2020.

10. Huang C, Wang Y, Li X, et al. Clinical features of patients infected with 2019 novel coronavirus in Wuhan, China. The Lancet 2020.

11. Wang D, Hu B, Hu C, et al. Clinical Characteristics of 138 Hospitalized Patients With 2019 Novel Coronavirus–Infected Pneumonia in Wuhan, China. JAMA 2020 doi: 10.1001/jama.2020.1585.

12. Rothe C, Schunk M, Sothmann P, et al. Transmission of 2019-nCoV infection from an asymptomatic contact in Germany. New England Journal of Medicine 2020

13. Update on the epidemic situation of new coronavirus pneumonia as of 24:00 on February 22, 2020. The National Health Commission of the People’s Republic of China

14. Novel Coronavirus(2019-nCoV) Situation Report - 7. World Health Organization 2020;https://www.who.int/docs/default-source/coronaviruse/situation-reports/20200127-sitrep-7-2019--ncov.pdf

15. Ferguson ND, Fan E, Camporota L, et al. The Berlin definition of ARDS: an expanded rationale, justification, and supplementary material. Intensive care medicine. 2012;38(10):1573–82.

16. Liu Y, Gayle AA, Wilder-Smith A, et al. The reproductive number of COVID-19 is higher compared to SARS coronavirus. Journal of Travel Medicine 2020.

17. WHO. Consensus document on the epidemiology of severe acute respiratory syndrome (SARS). 2003.

18. Yang X, Yu Y, Xu J, et al. Clinical course and outcomes of critically ill patients with SARS-CoV-2 pneumonia in Wuhan, China: a single-centered, retrospective, observational study. 2020.

19. Chu H, Zhou J, Wong BH-Y, et al. Middle East respiratory syndrome coronavirus efficiently infects human primary T lymphocytes and activates the extrinsic and intrinsic apoptosis pathways. The Journal of infectious diseases 2016;213(6):904–14.

20. Liu WJ, Zhao M, Liu K, et al. T-cell immunity of SARS-CoV: Implications for vaccine development against MERS-CoV. Antiviral research 2017;137:82–92.

21. Diao B, Wang C, Tan Y, et al. Reduction and Functional Exhaustion of T Cells in Patients with Coronavirus Disease 2019 (COVID-19). medRxiv 2020:2020.02.18.20024364. doi: 10.1101/2020.02.18.20024364.

22. Das KM, Lee EY, Jawder SEA, et al. Acute Middle East respiratory syndrome coronavirus: temporal lung changes observed on the chest radiographs of 55 patients. American Journal of Roentgenology 2015;205(3):W267–S74.

23. MÜller NL, Ooi GC, Khong PL, et al. High-resolution CT findings of severe acute respiratory syndrome at presentation and after admission. American Journal of Roentgenology 2004;182(1):39–44.

24. Nicolaou S, Al-Nakshabandi NA, MÜller NL. SARS: imaging of severe acute respiratory syndrome. American Journal of Roentgenology 2003;180(5):1247–49.

25. Gao H-N, Lu H-Z, Cao B, et al. Clinical findings in 111 cases of influenza A (H7N9) virus infection. New England Journal of Medicine 2013;368(24):2277–85.

26. Zhao Y, Zhao Z, Wang Y, et al. Single-cell RNA expression profiling of ACE2, the putative receptor of Wuhan 2019-nCov. BioRxiv 2020.

27. Zhou P, Yang X-L, Wang X-G, et al. A pneumonia outbreak associated with a new coronavirus of probable bat origin. Nature 2020:1–4.

28. Richardson P, Griffin I, Tucker C, et al. Baricitinib as potential treatment for 2019-nCoV acute respiratory disease. The Lancet 2020.

29. Holshue ML, DeBolt C, Lindquist S, et al. First case of 2019 novel coronavirus in the United States. New England Journal of Medicine 2020.

30. Wang M, Cao R, Zhang L, et al. Remdesivir and chloroquine effectively inhibit the recently emerged novel coronavirus (2019-nCoV) in vitro. Cell Research 2020:1–3.

31. Avaiable from: https://clinicaltrials.gov/ct2/results?cond=Coronavirus&term=&cntry=&state=&city=&dist=.

32. Russell CD, Millar JE, Baillie JK. Clinical evidence does not support corticosteroid treatment for 2019-nCoV lung injury. The Lancet 2020

33. Peter JV, John P, Graham PL, et al. Corticosteroids in the prevention and treatment of acute respiratory distress syndrome (ARDS) in adults: meta-analysis. Bmj 2008;336(7651):1006–09.

34. Khilnani GC, Hadda V. Corticosteroids and ARDS: A review of treatment and prevention evidence. Lung India 2011;28(2):114–19. doi: 10.4103/0970-2113.80324.

35. Roch A, Guervilly C, Papazian L. Fluid management in acute lung injury and ards. Ann Intensive Care 2011;1(1):16–16. doi: 10.1186/2110-5820-1-16.

